# Early transfusion of convalescent plasma improves the clinical outcome in severe SARS-CoV2 infection

**DOI:** 10.1101/2021.05.25.21257770

**Authors:** Eszter Fodor, Veronika Müller, Zsolt Iványi, Tímea Berki, Kuten Pella Olga, Mira Ambrus, Ágnes Sárkány, Árpád Skázel, Ágnes Madár, Dorottya Kardos, Gábor Kemenesi, Fanni Földes, Sándor Nagy, Andrea Matusovits, Nacsa János, Attila Tordai, Ferenc Jakab, Zsombor Lacza

## Abstract

Plasma harvested from convalescent COVID-19 patients (CCP) has been applied as first-line therapy in the early phase of the SARS-CoV2 pandemic through clinical studies using various protocols. We present data from a cohort of 267 hospitalized, severe COVID-19 patients who received CCP. No transfusion-related complications were reported, indicating the overall safety of CCP therapy. Patients who eventually died from COVID-19 received CCP significantly later (3.95 versus 5.22 days after hospital admission) and had higher interleukin 6 (IL-6) levels (28.9 pg/ml versus 102.5 pg/ml) than those who survived. In addition, CCP-transfusion caused a significant reduction in the overall inflammatory status of the patients regardless of the severity of disease or outcome, as evidenced by decreasing C-reactive protein, IL6 and ferritin levels. We conclude that, CCP-transfusion is a safe and effective supplementary treatment modality for hospitalized COVID-19 patients characterized by better expected outcome if applied as early as possible. We also observed that, IL-6 may be a suitable laboratory parameter for patient selection and monitoring of CCP therapy effectiveness.

## 1. Introduction

Severe acute respiratory syndrome caused by the novel coronavirus SARS-CoV2 has been responsible for an unprecedented world pandemic starting out from Vuhan (China) in January 2020. This represented an extraordinary challenge for the health care systems and entire societies of the developed countries [1,2] including Hungary [3]. The SARS-CoV2-induced COVID-19 disease is characterized by acute viral pneumonia associated with progressive respiratory insufficiency, requiring oxygen supplementation in about 15% and mechanical ventilation in 5% of the cases with reported overall mortality rates of 2.3% [4,5]. Since SARS-CoV2 is a new human pathogen, initially neither vaccines nor effective therapeutic modalities were available. With several earlier attempts of antiviral passive immunotherapy, COVID-19 convalescent plasma (CCP) therapy has quickly become a readily available and promising therapeutic modality against severe COVID-19 [6,7]. Given the large amount of clinical experience with allogeneic plasma transfusions and the relative lack of potential side effects, regulatory agencies have quickly granted permissions for several parallel clinical trials with CCP [8–10]. After CCP transfusion, virus neutralization antibodies represent the most important mode of action but moderating the unnecessarily severe immune reaction by several soluble factors has also been proposed [11–13].

Apart from the multitude of case studies, initial reports of CCP clinical studies on sizeable cohorts have not been able to demonstrate significant improvements in mortality rates [14–17]. In contrast, in subsequent studies, mortality-benefits were documented among unselected, severe COVID-19 patients after CCP-transfusions in the USA [18–21], in China [22,23], in India [24], in Europe [25,26] and among selected patients based on risk factors [27,28]. In addition, a potential benefit was also observed in a meta-analysis[29]. Development of severe COVID-19 associated respiratory disease was significantly less frequent after CCP transfusion administered within 72 hours of onset of mild COVID-19 infection compared to a randomized control group [30]. In an early meta-analyzes, low to moderate evidence indicated a lacking benefit [31], a later systematic compilation covering more than 35,000 CCP-treated cases indicated a significantly reduced odds ratio for all-cause mortality associated with CCP-transfusion [29]. Thus, outcome observations after CCP therapies are controversial due to the difficulties to form and analyze homogenous patient cohorts and control groups, the diverse availability of other therapeutic modalities and the substantial differences in local clinical practices. Most studies aimed at reducing mortality rates, however, this required a large number of recruited patients and homogenous treatment groups, which was seldom the case. However, quantitative laboratory parameters may serve as secondary endpoints and provide information on individual patients and also on potential patient selection criteria for future studies. Our earlier experience with blood plasma-based therapies has also provided evidence that measuring cytokine levels may be informative in monitoring the effectiveness of blood-based therapies [32,33]. Thus, the aim of the current study to analyze the outcome and immunology parameters after CCP-transfusions in severe COVID-19 patients.

## 2. Materials and Methods

### Study description

The trial was designed in accordance with WHO (2015 WHO/HIS/KER/GHE/15.1) as an open-label, prospective, interventional study and approved by the Hungarian National Medical Research Council (approval number: IV/3457/2-2020-EKU) and registered in ClinicalTrials.gov (NCT04345679). Due to ethical concerns of leaving any patient untreated or placebo-treated (e.g., non-convalescent fresh frozen plasma), no control arm was included. The primary clinical endpoint was patient survival, secondary endpoints were the time of convalescent plasma administration (days from hospital admission), the duration of hospital stay (days), the mortality rate (% of patients), the hematological laboratory parameter WBC, and inflammatory markers such as ferritin, C-reactive protein (CRP) and interleukin-6 (IL-6).

### Plasma donation

Plasma was collected from healthy voluntary convalescent donors of both sexes aged 18-65 years, with confirmed previous SARS-CoV2 infection. A positive result of either SARS-CoV2 PCR or antigen screening test at the time of plasma donation was a reason for exclusion. Donors were required to meet all eligibility requirements valid for regular blood donors. An inclusion criterion was a positive IgG antibody level (>24 U/ml) against SARS-CoV2 nucleocapsid protein (measured by Microgen semi-quantitative ELISA assays). To decrease the risks of TRALI (transfusion-related acute lung injury), all female donors were screened for anti-HLA antibodies, and positive donors were excluded from plasma donation. Altogether 400 ml of convalescent plasma was collected from donors through an automated plasma apheresis separator. The Hungarian National Blood Service divided the 400 ml plasma into two therapeutic doses of 200 ml. Plasma products underwent pathogen reduction and irradiation. The processed plasma was stored at -20°C until used. ABO compatibility was strictly observed throughout the study.

### Recipients

Convalescent plasma recipients were hospitalized patients of both sexes and ages admitted to a University hospital COVID ward between April 2020 and February 2021 and who were suitable for receiving plasma judged by their attending physicians.

### Laboratory measurements

Laboratory hematological parameters were measured with clinical hematology analyzers. Ferritin and C-reactive protein levels were measured before and on days 1 and 7 after plasma transfusion. White blood cell count was determined before and on days 1 and 2 after plasma transfusion. Levels of the pro-inflammatory cytokine interleukin-6 (IL-6) were measured before and on days 1 and 7 with electrochemiluminescence immunoassay (ECLIA).

### Simple Plex cytokine assay

IL-1 beta, IL-6, IL-8 and TNF alpha concentrations were analyzed in plasma from 21 selected COVID-19 patients with Simple Plex assays run on the Ella(tm) automated immunoassay analyzer (ProteinSimple, San Jose, CA). Diluted (1:2) plasma samples together with buffer were loaded into the ELLA cartridge and measured according to the manufacturer’s instruction.

### Statistical analysis

Data were analyzed with the two-tailed Mann-Whitney test and Fisher’s exact test using GraphPad Prism software. P<0.05 was considered significant.

## 3. Results

### 3.1 Patient characteristics

The key characteristics and laboratory findings of 267 patients can be found in Table 1. Data distribution was spread on a wide scale, demonstrated in histograms of Fig.1. The median of patients’ age was 67 years, with a median hospitalization time of 15 days. Over 90% of patients had serious co-morbities and had markedly elevated inflammatory parameters before CCP-treatment (Table 1). 76% of patients were admitted to the COVID ward (CW) and 24% to the ICU.

**Table.1.** Baseline characteristics of COVID-19 positive patients selected for CCP therapy. Patients’ parameters have been calculated for the whole group (n=267) and for COVID ward and ICU patients. Data were analyzed with Fisher’s exact test.

|  | All patients<br>n=267 | COVID<br>ward<br>(n=202) | ICU<br>(n=65) | P (< 0.05<br>significant) |
| --- | --- | --- | --- | --- |
| Age(years, median) | 67 | 69 | 63 | 0,0103 |
| Sex, n (%) | female 40% | female 45% | female 26% | 0,0089 |
|  | male 60% | male 55% | male 74% |  |
| Hospitalization time (days, median) | 15 | 14 | 17 | 0,0202 |
| Time until CCP therapy (days, median) | 3 | 3 | 3 | 0,8087, ns |
| Hypertension, n (%) | 63,5% | 49,5% | 49% | 0,2959, ns |
| Diabetes, n (%) | 25,5% | 22,3% | 12% | 0,1833, ns |
| Cancer active, n (%) | 11,5% | 10,4% | 5,3% | 0,3014, ns |
| Other chronic illness | 21,2% | 10,5% | 6,1% | 0,4626, ns |
| No chronic illness | 9,1% | 7,4% | 6,1% | 1, ns |
| Not known (no documentation about chronic illness) | 22,1% | 19% | 29% | - |
| Recovered, n (%) | 68,5% | 77,7% | 40% | P<0.0001 |
| Deceased, n (%) | 31,5% | 22,3% | 60% |  |
| WBC (G/l), mean (day 1) | 9,8 | 8,7 | 13,4 | 0,0010 |
| Ferritin (ng/ml), mean (day 1) | 1296 | 1026 | 2121 | 0,0003 |
| CRP (mg/L), mean (day 1) | 111,5 | 103,7 | 136,6 | 0,0090 |
| IL-6 (pg/ml), mean (day1) | 77,5 | 82.68 | 34.06 | 0,1452, ns |

|  | All patients<br>n=267 | COVID<br>ward<br>(n=202) | ICU<br>(n=65) | P (< 0.05<br>significant) |
| --- | --- | --- | --- | --- |
| Age(years, median) | 67 | 69 | 63 | 0,0103 |
| Sex, n (%) | female 40% | female<br>45% | female<br>26% | 0,0089 |
|  | male 60% | male 55% | male 74% |  |
| Hospitalization time (days,<br>median) | 15 | 14 | 17 | 0,0202 |
| Time until CCP therapy<br>(days, median) | 3 | 3 | 3 | 0,8087, ns |
| Hypertension, n (%) | 63,5% | 49,5% | 49% | 0,2959, ns |
| Diabetes, n (%) | 25,5% | 22,3% | 12% | 0,1833, ns |
| Cancer active, n (%) | 11,5% | 10,4% | 5,3% | 0,3014, ns |
| Other chronic illness | 21,2% | 10,5% | 6,1% | 0,4626, ns |
| No chronic illness | 9,1% | 7,4% | 6,1% | 1, ns |
| Not known (no<br>documentation about<br>chronic illness) | 22,1% | 19% | 29% | - |
| Recovered, n (%) | 68,5% | 77,7% | 40% | P<0.0001 |
| Deceased, n (%) | 31,5% | 22,3% | 60% |  |
| WBC (G/l), mean<br>(day 1) | 9,8 | 8,7 | 13,4 | 0,0010 |
| Ferritin (ng/ml), mean<br>(day 1) | 1296 | 1026 | 2121 | 0,0003 |
| CRP (mg/L), mean<br>(day 1) | 111,5 | 103,7 | 136,6 | 0,0090 |
| IL-6 (pg/ml), mean (day1) | 77,5 | 82.68 | 34.06 | 0,1452, ns |

**Fig. 1.**
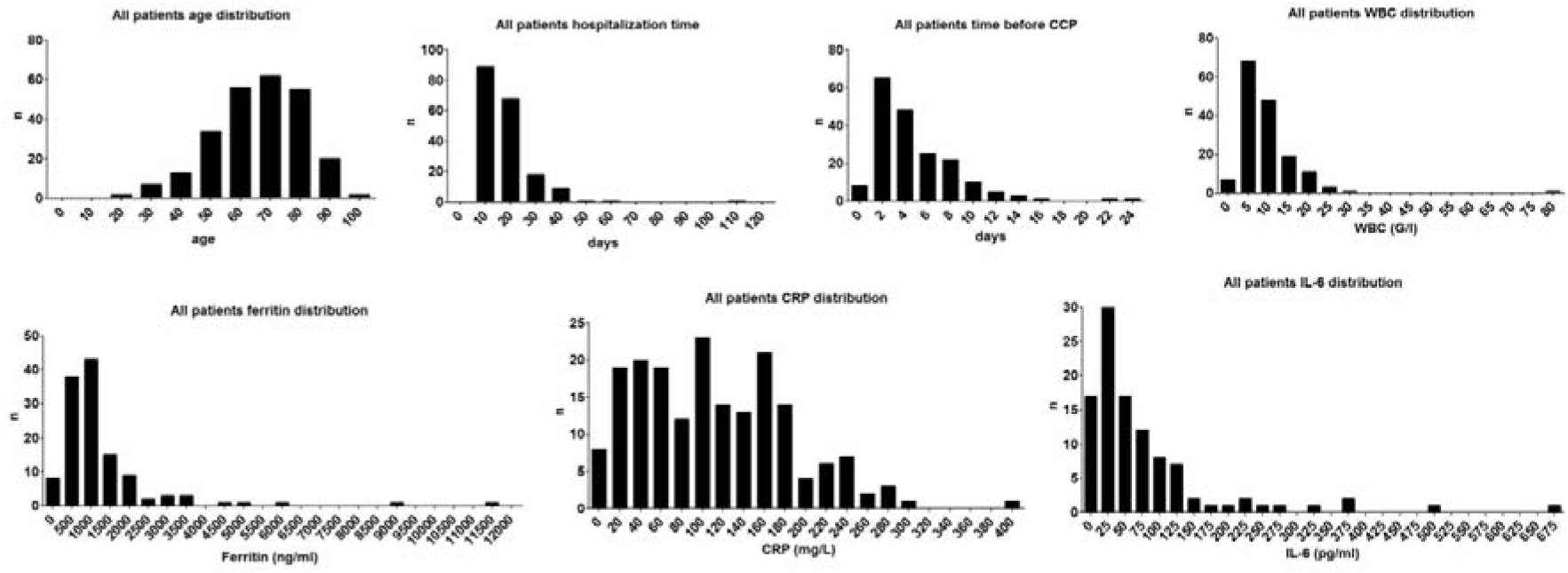
Histograms presenting the distribution of age, hospitalization time, time before CCP, and the levels of WBC, ferritin, CRP and IL-6 on day 1. The total number of patients analysed was 267, however, when laboratory tests were not performed on a given day data was excluded. Documentation about WBC level was available for n=158 patients, about ferritin for n=126 patients, about CRP for n=187 patients and about IL-6 for n=104 patients. Nonetheless, all statistical comparisons were adequately powered.

After comparing patients from CW and ICU a statistical relationship between age, sex, hospitalization time, mortality, WBC, ferritin and CRP level was found. The median age of patients admitted to the ICU was lower (63 years) than that of CW patients (69 years) and 74% of ICU patients were male. As expected, WBC, ferritin and CRP levels on the day of admission were significantly higher in the ICU patient group. Also, ICU patients had longer hospitalization time but the median number of days to CCP therapy was the same in both groups. No significant association with CW/ICU admission and the time until CCP therapy, chronic illness and IL-6 level was observed. Since the decision on whether a patient is treated at the ICU was based on multiple factors partly unrelated to their COVID-19 status, e.g., co-morbidities or place availability, and that there was no difference in the treatment regimens regarding plasma therapy or the inflammatory parameters between the CW and the ICU subgroups, the subsequent analyzes are presented as a single group of patients.

### 3.2 Administration of CCP to COVID patients

CCP transfusion was safe with no grade 3 adverse events reported. To identify the best time point when CCP should be administrated, the hospitalization time before CCP therapy was compared between the deceased and recovered patients (Fig.2). Frequency distribution analysis revealed that CCP therapy was applied earlier in patients who eventually recovered than in those who deceased, with a significant difference (P= 0,0133) between the time of CCP therapy between the survivors (3,95 days) and deceased patients (5,22 days) (Fig. 2). Based on this observation, we regrouped the patients to those who received CCP up to 3 days after hospitalization and compared their survival rate to those that received CCP at day 4 or later time points in their respective treatment regimes. Indeed, the mortality rate was significantly lower (P= 0,0226) among patients who received CCP up to 3 days after hospital admission (24%) versus patients who received it later (40%) (Fig. 2).

**Fig. 2.**
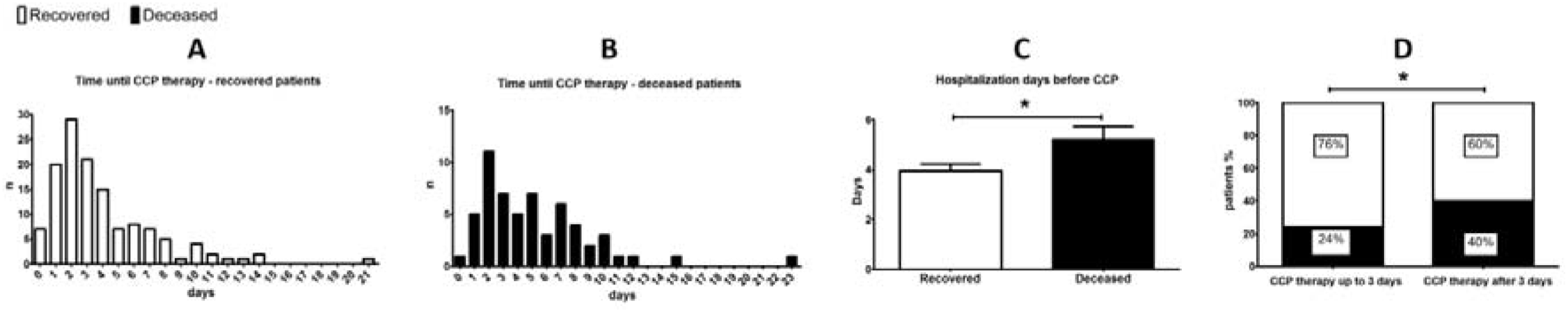
Time until CCP therapy in hospitalized patients. Panels A and B show histograms of time until CCP therapy in days from hospitalization in survivors and decedents, respectively. Panel C shows the mean ± SEM values of hospitalization time before CCP for recovered and deceased patients, asterisk indicates p<0.05 with Mann-Whitney test. Panel D shows that patients who received plasma during the first 3 days of hospitalization had a significantly better survival rate than those who received CCP therapy at later timepoints, asterisk indicates P<0.05 with Fisher’s exact test.

### 3.3 Effect of CCP therapy on inflammatory parameters

To identify the reliable prognostic factors for COVID patient’s recovery, IL-6, IL-1 beta, IL-8 and TNF alpha was additionally analyzed from the available frozen serum samples of 21 patients (Fig.3). IL-1 beta (deceased 0,5 pg/ml, recovered 0,6 pg/ml), IL-8 (deceased 31,3 pg/ml, recovered 22,5 pg/ml) and TNF alpha concentration (deceased 21,1 pg/ml, recovered 16,1 pg/ml), were not above the physiological level (minimum-maximum values of cytokines levels in healthy controls according to the literature are 0,0-5,0 pg/ml for IL-1 beta [34]; 0,0–50,4 pg/ml for IL-8 [35] and 0,0–32,5 pg/ml for TNF alpha [35]). Also, the changes of these cytokine levels were not significant between recovered and deceased patients (Fig. 3). At the same time, IL-6 concentration was clearly above the normal level for healthy individuals (0.0–12.7 pg/mL [35]) before plasma therapy and patients who died during hospitalization had significantly higher (P= 0,0066) IL-6 levels than survivors (deceased 102,5 pg/ml, recovered 28,9 pg/ml, Fig. 3).

**Fig. 3.**
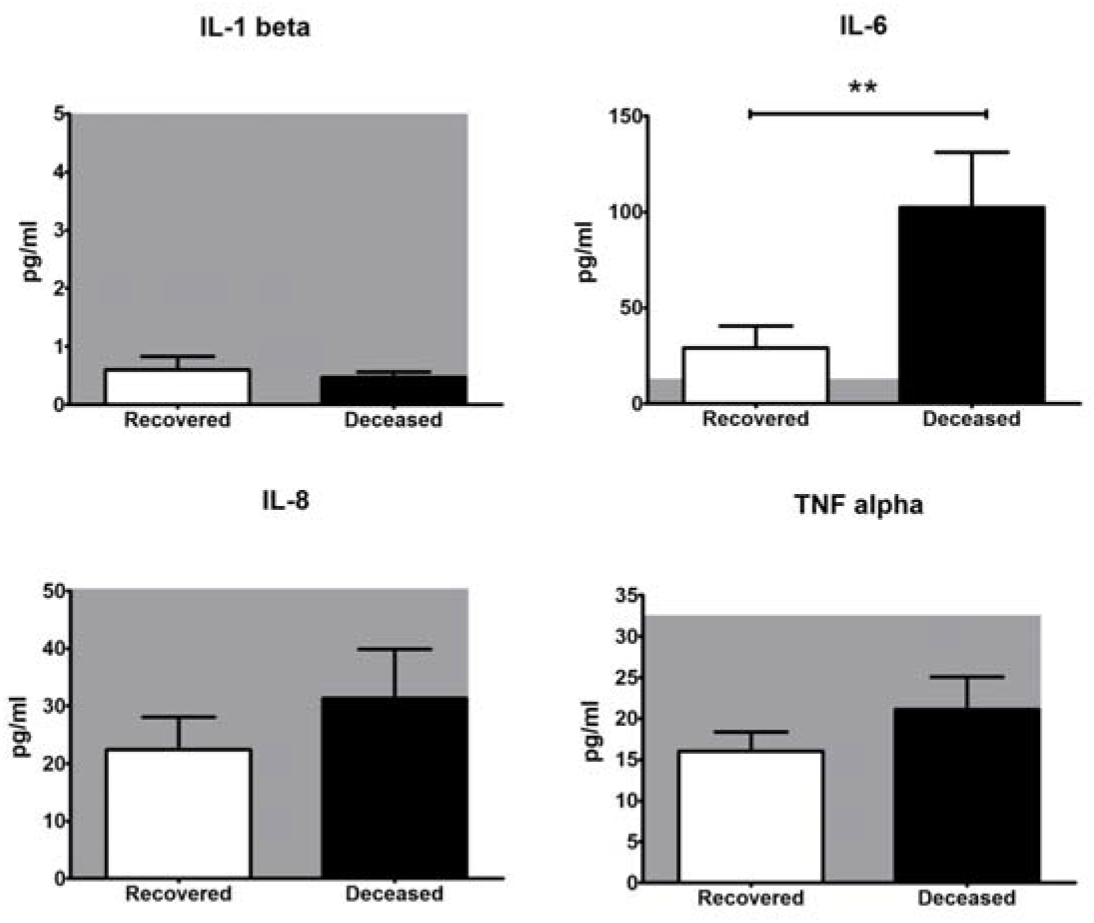
IL-6, IL-1 beta, IL-8 and TNF alpha levels in survivors and decedents blood harvested before CCP therapy (n=21 patients). Normal range is indicated with a greyed area on each panel. Data are represented as average ± SEM, asterisk indicates P<0.05 with Mann-Whitney’s test.

The level of ferritin, CRP and IL-6 was compared between blood samples collected before and after CCP therapy to assess if there was an improvement of patients' condition. All three parameters were significantly lower (ferritin P = 0,0025; CRP and IL-6 P<0.0001) after plasma administration (Fig. 4).

**Fig. 4.**
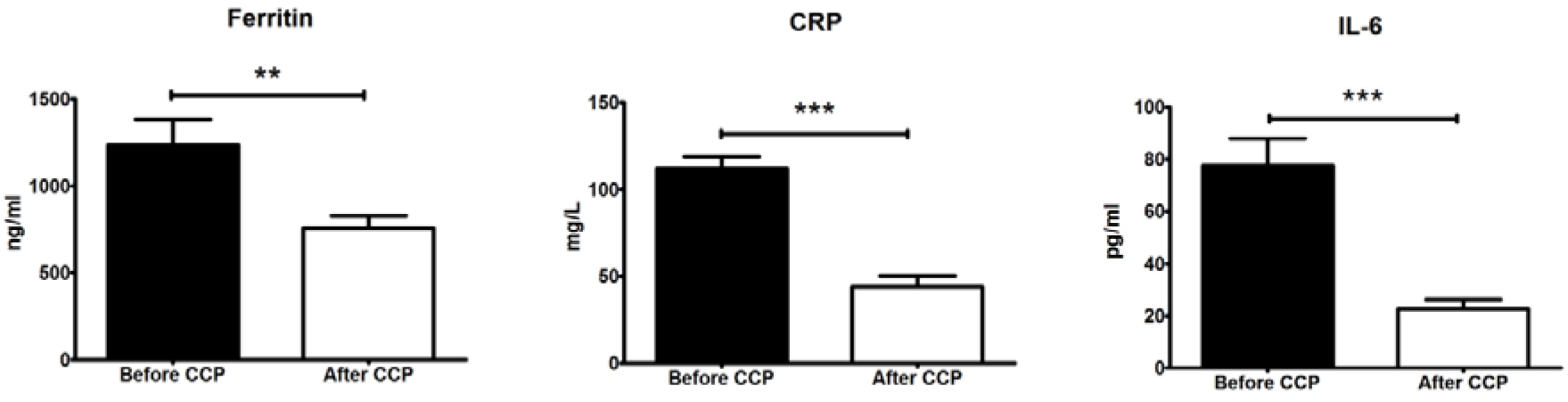
IL-6, CRP and ferritin levels before and after CCP. Documentation about the ferritin level before and after CCP was available for n= 79 patients, about CRP for n=130 patients and IL-6 for n=58 patients. Data are presented as average ± SEM, ** indicates P<0.01, *** indicates P<0.001 with Mann-Whitney’s test.

## 4. Discussion

In the current study, we systematically monitored selected outcome parameters in a sizeable cohort of severe COVID-19 patients who received CCP therapy as a supplementary intervention besides standard supportive and causative therapies. As expected, several baseline characteristics showed significant differences upon comparing those patients who were treated on the COVID ward versus those under intensive care reflecting the different severity of their overall clinical status. In agreement with other reports [24], males were over-represented in the ICU-subgroup, along with elevated WBC, ferritin and CRP levels. A high proportion of patients had severe co-morbidities that clearly affected their treatment needs in addition to their COVID19 status, therefore, it is not justified to separate the cohort into subgroups based on any variable singled out from a multitude of parameters.

We addressed the importance of initiation timepoint of CCP transfusions by using the indicator of number of days elapsing in the hospital prior to CCP therapy. In such comparison, significantly longer time elapsed in the deceased subgroup compared to survivors. Moreover, dichotomizing the entire patient group according to the median value of days until CCP-therapy with 3 days as a threshold, showed significantly higher proportion of deceased patients in the subgroup characterized by longer than 3 days. Supporting the importance of early application of CCP therapy comes from a prospective USA study indicating significant mortality reduction compared to matched control cases only in the subgroup receiving CCP transfusion within 72 hours of hospital admission [20]. In a smaller comparative study CCP-transfusion within 72 hours from hospital admission was more efficient among patients aged less than 65 years [19]. In another smaller study a similar benefit of earlier application of CCP-therapy was observed with improvements of several clinical outcomes associated with CCP-administration before 7 days from diagnosis [26]. Furthermore, in an outstanding randomized, double-blind, placebo-controlled trial performed in Argentina, development of COVID-19-associated predefined severe respiratory disease was significantly less frequent in the treated group receiving CCP transfusion within 72 hours from the onset of mild COVID-19 in an elderly population [30]. The importance of appropriate timing of CCP-transfusions as well as careful candidate selection instead of the compassionate use of CCP has clearly been emphasized in an editorial [36].

Considering the relative paucity of information on inflammatory cytokine levels in CCP-treated COVID-19-patients, in a subgroup of our cohort, we included baseline and follow-up measurements of critical cytokine levels such as IL-6, IL-8, TNF-alpha and IL-1-beta. Comparing baseline levels between recovered and deceased patients, we observed that IL-6 was the only inflammatory cytokine characterized by significantly elevated baseline levels among the deceased CCP-treated patients indicating a potential role of this inflammatory cytokine as a prognostic marker. In another comparison addressing marker status before and after CCP-transfusion, similarly to general inflammatory markers, IL-6 levels showed significant decreases. This observation is in line with those of others and suggests a putative immunomodulatory effect exerted by CCP [11]. In particular, in smaller study, besides IL-6, additionally TNF-alpha and IFN-gamma levels also showed significant decreases while that of IL-10 increased following CCP-therapy [37]. In the framework of another randomized trial of CCP therapy, Il-6 serum levels decreased along with those of interferon gamma induced protein 10 in response to CCP-transfusion [38]. However, it should be noted that, the possible connection of IL-6 levels with the therapeutic effect of CCP does not mean a causal relationship in either way, and further studies are needed to identify the molecular-level interactions between IL-6-involving inflammatory mechanisms. It is also noteworthy that, convalescent plasma is also a source of a number of interleukins and cytokines that are administered alongside the anti-SARS2-CoV neutralizing antibodies.

Limitations of the current study include its non-controlled design, its limited sample size and partial unavailability of samples for detailed immunological testing in a significant proportion of the patients included. Pre-hospital history of the patients was not accessible leaving room for variation in SARS2-CoV2 infection periods. In light of all the above limitations, we believe that our observations should rather be viewed as an indicator prompting further studies with improved design targeting special features of CCP therapy such as early intervention in patients with only moderate IL-6 levels. Comparing results of the large number of registered clinical trials (n=164 on ClinicalTrials.gov at the time of writing) may help to identify critical factors in study design and patient selection to obtain benefits of clinical outcome after CCP in severe COVID-19, however, this future effort will depend on original study data to become available.

In conclusion, our results indicate that CCP transfusion is a safe supplementary option that can easily be incorporated into the complex therapeutic scheme of severe COVID-19 patients. Early timing of CCP transfusion is clearly preferable. CCP transfusion resulted in significant decreases in inflammatory markers and pretransfusion measurements of these markers may allow further stratification of severe COVID-19 patients amenable for efficient CCP therapy.

## Data Availability

Due to the nature of this research, participants of this study did not agree for their data to be shared publicly, so supporting data is not available

## Author Contributions

Conceptualization, Zsombor Lacza and Eszter Fodor and Olga Kuten.; methodology, Veronika Müller, validation, Ferenc Jakab, Gábor Kemenesi, Fanni Földes, formal analysis, Ágnes Madár, Mira Ambrus and Dorottya Kardos; investigation, Zsolt Iványi.; data curation, Eszter Fodor and Attila Tordai, János Nacsa; writing— original draft preparation, Zsombor Lacza, Olga Kuten; writing—review and editing, supervision, Attila Tordai, Zsombor Lacza, Eszter Fodor, Tímea Berki; project administration, Árpád Skázel, resources, Sándor Nagy, Andrea Matusovits, Ágnes Sárkány

## Funding

This research was funded by the Ministry for Innovation and Technology and the grant numbers are the following: K135757 and 2020-1.1.6-JÖVŐ-2021-00010. 4

## Institutional Review Board Statement

The study was conducted according to the guidelines of the Declaration of Helsinki, and approved by the Hungarian National Ethics Comittee (protocol code 1943-6/2020/EÜIG and date of approval: 2020.04.15).

## Informed Consent Statement

Informed consent was obtained from all subjects involved in the study.

## Acknowledgments

This work was partially supported by the Higher Education Institutional Excellence Program (FIKP) of the Ministry for Innovation and Technology, within the framework of the Molecular Biology thematic program of the Semmelweis University as well as by grants from the National Research, Development and Innovation Fund (NKFI) K135757 and 2020-1.1.6-JÖVŐ-2021-00010. We would like to thank you all the donors, who donated convalescent plasma, and all the nurses, doctors and health care workers who made this treatment possible.

## Conflicts of Interest

The authors declare no conflict of interest. Zsombor Lacza owns stock in OrthoSera kft, a startup company developing the hyperacute serum technology towards clinical application.

## Notes

### Competing Interest Statement

The authors have declared no competing interest.

### Clinical Trial

NCT04345679

### Author Declarations

The study was conducted according to the guidelines of the Declaration of Helsinki, and approved by the Hungarian National Ethics Comittee (protocol code 1943-6/2020/EUIG and date of approval: 2020.04.15).

## References

1. Guan W, Ni Z, Hu Y, Liang W, Ou C, He J, et al. Clinical Characteristics of Coronavirus Disease 2019 in China. N Engl J Med. 2020 Apr 30;382(18):1708–20.

2. Richardson S, Hirsch JS, Narasimhan M, Crawford JM, McGinn T, Davidson KW, et al. Presenting Characteristics, Comorbidities, and Outcomes among 5700 Patients Hospitalized with COVID-19 in the New York City Area. JAMA - J Am Med Assoc. 2020 May 26;323(20):2052–9.

3. Tordai A, Nagy S, Baróti-Tóth K, Marton I, Lázár M, Demeter J, et al. A SARS-CoV2-járvány hatása a hazai vérellátásra. Hematológia–Transzfuziológia. 2020 Sep 12;53(2):96–105.

4. Chen N, Zhou M, Dong X, Qu J, Gong F, Han Y, et al. Epidemiological and clinical characteristics of 99 cases of 2019 novel coronavirus pneumonia in Wuhan, China: a descriptive study. Lancet. 2020 Feb 15;395(10223):507–13.

5. Wu Z, McGoogan JM. Characteristics of and Important Lessons from the Coronavirus Disease 2019 (COVID-19) Outbreak in China: Summary of a Report of 72314 Cases from the Chinese Center for Disease Control and Prevention. Vol. 323, JAMA - Journal of the American Medical Association. American Medical Association; 2020. p. 1239–42.

6. Casadevall A, Pirofski LA. The convalescent sera option for containing COVID-19 [Internet]. Vol. 130, Journal of Clinical Investigation. American Society for Clinical Investigation; 2020. p. 1545–8.

7. Abraham J. Passive antibody therapy in COVID-19. Vol. 20, Nature Reviews Immunology. Nature Research; 2020. p. 401–3.

8. Recommendations for Investigational COVID-19 Convalescent Plasma | FDA [Internet].

9. Tanne JH. Covid-19: FDA approves use of convalescent plasma to treat critically ill patients. BMJ. 2020 Mar 26;368:m1256.

10. Epstein J, Burnouf T. Points to consider in the preparation and transfusion of COVID-19 convalescent plasma. Vol. 115, Vox Sanguinis. Blackwell Publishing Ltd; 2020. p. 485–7.

11. Rojas M, Rodríguez Y, Monsalve DM, Acosta-Ampudia Y, Camacho B, Gallo JE, et al. Convalescent plasma in Covid-19: Possible mechanisms of action. Vol. 19, Autoimmunity Reviews. Elsevier B.V.; 2020.

12. Simon M, Major B, Vácz G, Kuten O, Hornyák I, Hinsenkamp A, et al. The effects of hyperacute serum on the elements of the human subchondral bone marrow niche. Stem Cells Int. 2018;2018.

13. Kardos D, Marschall B, Simon M, Hornyák I, Hinsenkamp A, Kuten O, et al. Investigation of Cytokine Changes in Osteoarthritic Knee Joint Tissues in Response to Hyperacute Serum Treatment. Cells. 2019 Aug 3;8(8).

14. Li L, Zhang W, Hu Y, Tong X, Zheng S, Yang J, et al. Effect of Convalescent Plasma Therapy on Time to Clinical Improvement in Patients with Severe and Life-threatening COVID-19: A Randomized Clinical Trial. JAMA - J Am Med Assoc. 2020 Aug 4;324(5):460–70.

15. Agarwal A, Mukherjee A, Kumar G, Chatterjee P, Bhatnagar T, Malhotra P. Convalescent plasma in the management of moderate covid-19 in adults in India: Open label phase II multicentre randomised controlled trial (PLACID Trial). BMJ. 2020 Oct 22;371.

16. Sostin O V., Rajapakse P, Cruser B, Wakefield D, Cruser D, Petrini J. A matched cohort study of convalescent plasma therapy for COVID-19. J Clin Apher. 2021;

17. Simonovich VA, Burgos Pratx LD, Scibona P, Beruto M V., Vallone MG, Vázquez C, et al. A Randomized Trial of Convalescent Plasma in Covid-19 Severe Pneumonia. N Engl J Med. 2021 Feb 18;384(7):619–29.

18. Shenoy AG, Hettinger AZ, Fernandez SJ, Blumenthal J, Baez V. Early mortality benefit with COVID-19 convalescent plasma: a matched control study. Br J Haematol. 2021 Feb 1;192(4):706–13.

19. Yoon H, Bartash R, Gendlina I, Rivera J, Nakouzi A, Bortz RH, et al. Treatment of severe COVID-19 with convalescent plasma in Bronx, NYC. JCI Insight. 2021 Feb 22;6(4).

20. Salazar E, Christensen PA, Graviss EA, Nguyen DT, Castillo B, Chen J, et al. Treatment of Coronavirus Disease 2019 Patients with Convalescent Plasma Reveals a Signal of Significantly Decreased Mortality. Am J Pathol. 2020 Nov 1;190(11):2290–303.

21. Joyner MJ, Senefeld JW, Klassen SA, Mills JR, Johnson PW, Theel ES, et al. Effect of convalescent plasma on mortality among hospitalized patients with COVID-19: Initial three-month experience. medRxiv; 2020. p. 2020.08.12.20169359.

22. Duan K, Liu B, Li C, Zhang H, Yu T, Qu J, et al. Effectiveness of convalescent plasma therapy in severe COVID-19 patients. Proc Natl Acad Sci U S A. 2020 Apr 28;117(17):9490–6.

23. Xia X, Li K, Wu L, Wang Z, Zhu M, Huang B, et al. Improved clinical symptoms and mortality among patients with severe or critical COVID-19 after convalescent plasma transfusion. Vol. 136, Blood. American Society of Hematology; 2020. p. 755–9.

24. Budhiraja S, Dewan A, Aggarwal R, Singh O, Juneja D, Pathak S, et al. Effectiveness of convalescent plasma in Indian patients with COVID-19. Blood Cells, Mol Dis. 2021 May 1;88.

25. Tworek A, Jaroń K, Uszyńska-Kałuża B, Rydzewski A, Gil R, Deptała A, et al. Convalescent plasma treatment is associated with lower mortality and better outcomes in high-risk COVID-19 patients – propensity-score matched case-control study. Int J Infect Dis. 2021 Apr 1;105:209–15.

26. Moniuszko-Malinowska A, Czupryna P, Zarębska-Michaluk D, Tomasiewicz K, Pancewicz S, Rorat M, et al. Convalescent Plasma Transfusion for the Treatment of COVID-19—Experience from Poland: A Multicenter Study. J Clin Med. 2020 Dec 24;10(1):28.

27. Liu STH, Lin HM, Baine I, Wajnberg A, Gumprecht JP, Rahman F, et al. Convalescent plasma treatment of severe COVID-19: a propensity score–matched control study. Nat Med. 2020 Nov 1;26(11):1708–13.

28. Pappa V, Bouchla A, Terpos E, Thomopoulos TP, Rosati M, Stellas D, et al. A phase ii study on the use of convalescent plasma for the treatment of severe covid-19-a propensity score-matched control analysis. Microorganisms. 2021 Apr 1;9(4).

29. Klassen SA, Senefeld JW, Johnson PW, Carter RE, Wiggins CC, Shoham S, et al. The Effect of Convalescent Plasma Therapy on COVID-19 Patient Mortality: Systematic Review and Meta-analysis. Mayo Clin Proc. 2021 May;96(5).

30. Libster R, Pérez Marc G, Wappner D, Coviello S, Bianchi A, Braem V, et al. Early High-Titer Plasma Therapy to Prevent Severe Covid-19 in Older Adults. N Engl J Med. 2021 Feb 18;384(7):610–8.

31. Janiaud P, Axfors C, Schmitt AM, Gloy V, Ebrahimi F, Hepprich M, et al. Association of Convalescent Plasma Treatment with Clinical Outcomes in Patients with COVID-19: A Systematic Review and Meta-analysis. JAMA - J Am Med Assoc. 2021 Mar 23;325(12):1185–95.

32. Kardos D, Marschall B, Simon M, Hornyák I, Hinsenkamp A, Kuten O, et al. Investigation of Cytokine Changes in Osteoarthritic Knee Joint Tissues in Response to Hyperacute Serum Treatment. Cells 2019, Vol 8, Page 824. 2019 Aug;8(8):824.

33. Neubauer M, Kuten O, Stotter C, Kramer K, Luna A De, Muellner T, et al. The Effect of Blood-Derived Products on the Chondrogenic and Osteogenic Differentiation Potential of Adipose-Derived Mesenchymal Stem Cells Originated from Three Different Locations. Stem Cells Int. 2019;2019.

34. Iglesias Molli AE, Bergonzi MF, Spalvieri MP, Linari MA, Frechtel GD, Cerrone GE. Relationship between the IL-1β serum concentration, mRNA levels and rs16944 genotype in the hyperglycemic normalization of T2D patients. Sci Rep. 2020 Dec 1;10(1).

35. Arican O, Aral M, Sasmaz S, Ciragil P. Serum levels of TNF-α, IFN-γ, IL-6, IL-8, IL-12, IL-17, and IL-18 in patients with active psoriasis and correlation with disease severity. Mediators Inflamm. 2005 Oct 24;2005(5):273–9.

36. Katz LM. (A Little) Clarity on Convalescent Plasma for Covid-19. N Engl J Med. 2021 Feb 18;384(7):666–8.

37. Pouladzadeh M, Safdarian M, Eshghi P, Abolghasemi H, bavani AG, Sheibani B, et al. A randomized clinical trial evaluating the immunomodulatory effect of convalescent plasma on COVID-19-related cytokine storm. Intern Emerg Med. 2021 Apr 10;

38. Bandopadhyay P, Lahiri A, Sarif J, Ray Y, Ranjan Paul S, Roy R, et al. Nature and dimensions of the systemic hyper-inflammation and its attenuation by convalescent plasma in severe COVID-19.

